# Effectiveness of inhaled therapies in asthma among adults in Northern Sri Lanka, a low and middle-income country: A prospective observational study

**DOI:** 10.1101/2024.07.08.24309593

**Authors:** Yalini Guruparan, Thiyahiny S Navaratinaraja, Gowry Selvaratnam, Shalini Sri Ranganathan

## Abstract

**Background:** Currently inhaled corticosteroids (ICS) alone, or in combined with inhaled long- acting beta_2_-agonist (LABA) is recommended for chronic asthma.

**Objective:** This study aimed to assess the effectiveness of inhaled therapies in a cohort of adult patients with asthma who were receiving treatment in a tertiary hospital in the Northern Sri Lanka.

**Methods:** A prospective cohort study was carried out among adult patients with asthma on inhaled medications for at least three months. Participants were followed up for six months with two follow-up interviews three months apart. Primary outcome measure was asthma control which was assessed by a locally validated asthma control patient-reported outcome measure (AC-PROM). Secondary outcome measures were use of short-acting beta_2_-agonists, number of nebulisations and number of hospitalisations. Chi-squared test was used to determine the significance of differences in outcome measures between the two groups. Logistic regression was performed to determine the association between asthma control and socio-demographic factors. A p-value ≤ 0.05 was considered statistically significant.

**Results:** Data from 1094 participants were analysed. Majority were females (73%) and belonged to age group >60 years (60%). Ratio between ICS monotherapy and combined therapy with ICS and LABA (ICS/LABA) was 3:1. A progressive improvement in asthma control was observed in both groups which was significant in those on ICS monotherapy (p<0.001). A significant reduction was also observed in overuse of short-acting beta_2_-agonist (p<0.001) and number of nebulisations (p=0.027) in participants on ICS monotherapy. No significant association between asthma control and socio-demographic factors was found in either group.

**Conclusions:** Both ICS monotherapy and ICS/LABA were effective. However, treatment package comprising regular ICS plus non-pharmacological approaches would be more realistic and cost-effective treatment strategy in the local context. Considering the low availability and current economic status of Sri Lanka, ICS/LABA could be reserved for poorly controlled asthma.

**What is already known on this topic?:** Inhaled corticosteroids (ICS) alone, or in combined with inhaled long-acting beta_2_-agonist (LABA) are recommended for the treatment of asthma. However, availability and affordability limit the use of inhaled medications in low and middle-income countries, particularly the combined therapy with ICS and LABA (ICS/LABA).

**What this study adds:** This study was conducted in a real-life clinical setting to compare the effectiveness of ICS monotherapy and ICS/LABA. We found that close monitoring and improved communication positively impacted asthma control and exacerbations in both ICS monotherapy and ICS/LABA. These changes were significant in those on ICS monotherapy, but not in those on ICS/LABA.

**How this study might affect research, practice, or policy:** Our study highlighted the importance of non-pharmacological approaches in the management of asthma. Promoting non-pharmacological approaches in routine clinical practice is likely to improve asthma control and reduce the need for escalation of treatment leading to improved quality of life and reduced healthcare costs.

## Introduction

Asthma is one of the common global health problems affecting all age groups and the prevalence of asthma is rising worldwide.[1] It was reported that in Sri Lanka prevalence of asthma in adults was 11%.[2] Though asthma is not curable, it is controllable with appropriate treatment.[3] Despite the availability of effective therapies and asthma treatment guidelines, more than half of the patients experience uncontrolled asthma.[4-5] Uncontrolled asthma increases the risk of exacerbations and economic burden to families and health care system of the country.[6-8]

Currently, inhaled corticosteroids (ICS) alone, or with inhaled long-acting beta_2_-agonist (LABA) is recommended as the first-line treatment of asthma.[1] However, in low and middle-income countries (LMICs) the availability and affordability of inhaled medications for asthma is poor.[9-11] Universal access to effective treatment is the key element in the management of non-communicable diseases (NCDs) including asthma. Therefore, the World Health Organization has set a target of 80% availability of medications required to treat major NCDs by 2025. [12] Poor availability and affordability of inhaled medications hinder effective asthma control in LMICs.[9-11] It is an urgent need to ensure universal access to essential asthma medicines in LMICs.

Sri Lanka is a LMIC with majority of the people relying on free public sector healthcare services.[13-14] However, a significant proportion of healthcare expenditure is from out-of-pocket including purchasing medications from private pharmacies.[15] In the public sector, availability of ICS monotherapy was reported to be 81% meeting the WHO’s target whereas the availability of combined therapy with ICS and LABA (ICS/LABA) was very low (17%). In the private sector availability of ICS monotherapy and ICS/LABA were 75% and 63% respectively.[16] Price of ICS/LABA was two and half times higher than ICS monotherapy which could account for this disparity.[17] Lack of availability in the public sector and unaffordability in the private sector could have limited the use of ICS/LABA in the management of asthma.

Many studies have been conducted to assess the effectiveness of different treatments for asthma including inhaled therapies in high-income countries.[18-20] However, such studies are limited in LMICs. Our study aimed to assess the effectiveness of inhaled therapies (ICS monotherapy versus ICS/LABA) in asthma control in a cohort of adult patients who were receiving treatment at Teaching Hospital-Jaffna, a tertiary hospital in the Northern Sri Lanka using a locally validated asthma control patient-reported outcome measure (AC-PROM). [21]

## Methods

This was a prospective cohort study among adult patients with asthma at Teaching Hospital, Jaffna which is the largest tertiary care hospital in the Northern Province of Sri Lanka and offers primary, secondary and tertiary health care services to the people residing in Jaffna district.

Adult patients with asthma on inhaled medication for at least three months were included in this study. Those with chronic obstructive pulmonary disease and tuberculosis were excluded. Sample size per group to determine the statistically significant difference in asthma control between ICS monotherapy and ICS/LABA was estimated with 90% power [22] using the proportion of patients with controlled asthma in ICS monotherapy (49%) and ICS/LABA (63%) reported by Pauwls *et al*.[23]

### Case definition of asthma

In this study asthma was defined as “symptoms such as wheeze, shortness of breath, cough, and chest tightness that vary over time and intensity together with variable airflow limitation”.[1]

### Primary and secondary outcome measures

The primary outcome measure for the effectiveness of inhaled therapies was asthma control which was determined by AC-PROM score.[21]

Secondary outcome measures include frequency of usage of short-acting beta_2_-agonist (SABA), number of nebulisations and number of hospitalisations due to exacerbation of asthma.

### Study instrument

A pretested interviewer-administered questionnaire was used for data collection. The questionnaire had three parts namely baseline, first follow-up and second follow-up. Baseline questionnaire sought information on socio-demographic characteristics, medication history, asthma care history, usage of SABA, number of nebulisations, number of hospital admissions and adherence. Follow-up questionnaires contained the same sections as the baseline questionnaire except the socio-demographic characteristics. All three parts contained AC-PROM to assess asthma control at the time of recruitment and first and second follow-ups. The AC-PROM was a locally developed tool which was validated against forced expiratory volume in 1 second (FEV_1_) of patients with asthma. It contains eight items assessing symptoms (4 items), exacerbation (2 items) and limitation of activity (2 items).[21]

### Recruitment and follow-up

Around 2500 adult patients with asthma were being followed up in medical clinics. These patients were screened for eligibility using a pre-recruitment checklist. Those on inhaled medications for at least three months were recruited into the cohort consecutively from December 2019 to June 2020. Each participant was followed up for six months with two follow-up interviews at three-month intervals. The first author personally collected the data.

Informed written consent was obtained from each participant before recruitment. Ethical approval was obtained from the Ethics Review Committee, Faculty of Medicine, University of Colombo, Sri Lanka (EC-18-108) and administrative approvals were obtained from relevant authorities before commencing the data collection.

### Data analysis

Descriptive statistics such as frequency, percentage, mean and standard deviation (SD were used to present the results. Asthma control was determined using the AC-PROM score. The cut-off value of the AC-PROM score for asthma control was ≥28.5. [21 Guruparan] Use of SABA four, or more than four times per week was considered as overuse indicating inadequate control of asthma. [24] Monthly household income was categorised based on the latest (2016) Sri Lankan household income.[25] Percent change was used to measure the changes in primary and secondary outcome measures between base-line and second follow-up. Chi-squared test was performed to determine the significance of changes in the outcome measures. Logistic regression was performed to determine the association of socio-demographic factors with asthma control. A p value ≤ 0.05 was considered statistically significant.

## Results

A total of 1200 participants were recruited into the cohort. Of them 1118 completed the two follow-ups (completion rate was 93.2%). Participants on SABA alone (n=2) and those who were taking oral medications in addition to inhaled therapy (n=22) were excluded from the analysis. Data from 1094 participants were analysed. Out of 1094 participants three fourth (75.6%; n=827) were on ICS monotherapy (beclomethasone dipropionate) and one-fourth (24.4%; n=267) were on ICS/LABA (fluticasone propionate/salmeterol=245 and budesonide/formoterol=22). During the follow-up no changes were observed in the treatment of asthma in any of the participants in either group.

Table 1 shows the socio-demographic characteristics of the participants. Mean age of the participants was 61.2 years (SD±11.6) and majority (60%) belonged to the age group >60 years. Around three-quarters of the participants (73%) were females. Most of the participants (60%) were housewives. More than half (57.6%) had low monthly household income. None of the participants in either group were current smokers. There was no significant association between asthma control and socio-demographic factors in either group.

**Table 1.** Socio-demographic characteristics of the participants.

| **Variables** | **Total population**<br>**(n=1094)** | **ICS monotherapy**<br>**(n=827)** | **ICS/LABA**<br>**(n=267)** |
| --- | --- | --- | --- |
| Age in years |  |  |  |
| ≤ 40 | 65 (5.9%) | 53 (6.4%) | 12 (4.5%) |
| 41-60 | 375 (34.3%) | 288 (34.8%) | 87 (32.6%) |
| > 60 | 654 (59.8%) | 486 (58.8%) | 168 (62.9%) |
| Gender |  |  |  |
| Male | 300 (27.4%) | 221 (26.7%) | 79 (29.6%) |
| Female | 794 (72.6%) | 606 (73.3%) | 188 (70.4%) |
| Educational level |  |  |  |
| Primary | 306 (28%) | 216 (26.1%) | 90 (33.7%) |
| Secondary | 655 (59.9%) | 515 (62.3%) | 140 (52.4%) |
| Higher | 133 (12.1%) | 96 (11.6%) | 37 (13.9%) |
| Employment status |  |  |  |
| Employed | 254 (23.2%) | 185 (22.4%) | 69 (25.8%) |
| Housewife | 656 (60%) | 537 (64.9%) | 119 (44.6%) |
| Pensioner | 63 (5.7%) | 36 (4.4%) | 27 (10.1%) |
| Unemployed | 121 (11.1%) | 69 (8.3%) | 52 (19.5%) |
| Monthly household income |  |  |  |
| Low | 630 (57.6%) | 521 (63%) | 109 (40.8%) |
| Middle | 260 (23.8%) | 176 (21.3%) | 84 (31.5%) |
| High | 133 (12.1%) | 130 (15.7%) | 74 (27.7%) |
| Smoking History |  |  |  |
| Smokers | 0 (0%) | 0 (0%) | 0 (0%) |
| Ex-smokers | 62 (5.7%) | 45 (5.4%) | 17 (6.4%) |
| Non-smokers | 1032 (94.3%) | 782 (94.6%) | 250 (93.6%) |

Table 2 shows the primary and secondary outcome measures. A progressive improvement in asthma control was observed in both groups. However, the improvement was significant only in those on ICS monotherapy (p<0.0001).

**Table 2.** Primary and secondary outcome measures over a period of six months.

| **Variables** | **Total**<br>**(n=1094)** | **ICS monotherapy**<br>**(n=827)** | **ICS/LABA**<br>**(n=267)** |
| --- | --- | --- | --- |
| **Asthma control** |  |  |  |
| Baseline | 653 (59.7%) | 450 (54.4%) | 203 (76%) |
| First follow-up | 759 (69.4%) | 547 (66.1%) | 212 (79.4%) |
| Second follow-up | 809 (73.9%)* | 593 (71.7%)* | 216 (80.9%) |
| **Overuse of SABA** |  |  |  |
| Baseline | 485 (44.3%) | 410 (49.6%) | 75 (28.1%) |
| First follow-up | 395 (36.1%) | 330 (39.9%) | 65 (24.3%) |
| Second follow-up | 360 (32.9%)* | 296 (35.8%)* | 64 (24%) |
| **Nebulisation** |  |  |  |
| Baseline | 121 (11.1%) | 93 (11.2%) | 28 (10.4%) |
| First follow-up | 92 (8.4%) | 68 (8.2%) | 24 (9%) |
| Second follow-up | 87 (7.9%)* | 64 (7.7%)* | 23 (8.6%) |
| **Hospitalisation** |  |  |  |
| Baseline | 22 (2%) | 15 (1.8%) | 7 (2.6%) |
| First follow-up | 18 (1.6%) | 12 (1.4%) | 6 (2.2%) |
| Second follow-up | 11 (1%) | 6 (0.7%) | 5 (1.9%) |

ICS- inhaled corticosteroid, LABA- long-acting beta2-agonist, \*p-value <0.05

Though all the secondary outcome measures showed a progressive improvement in both groups, statistically significant reduction was observed in overuse of SABA (p<0.001) and nebulisation (p=0.02) in those on ICS monotherapy (Table 2).

Percent changes in asthma control, overuse of SABA, number of nebulisations and number of hospitalisations when comparing baseline and second follow-up are shown in Figure 1. Almost five times greater improvement in asthma control was observed in participants on ICS monotherapy compared to those on ICS/LABA. Reduction in overuse of SABA, nebulisation and hospitalisation among patients receiving ICS monotherapy was almost twice as much as those on ICS/LABA.

**Figure 1.**
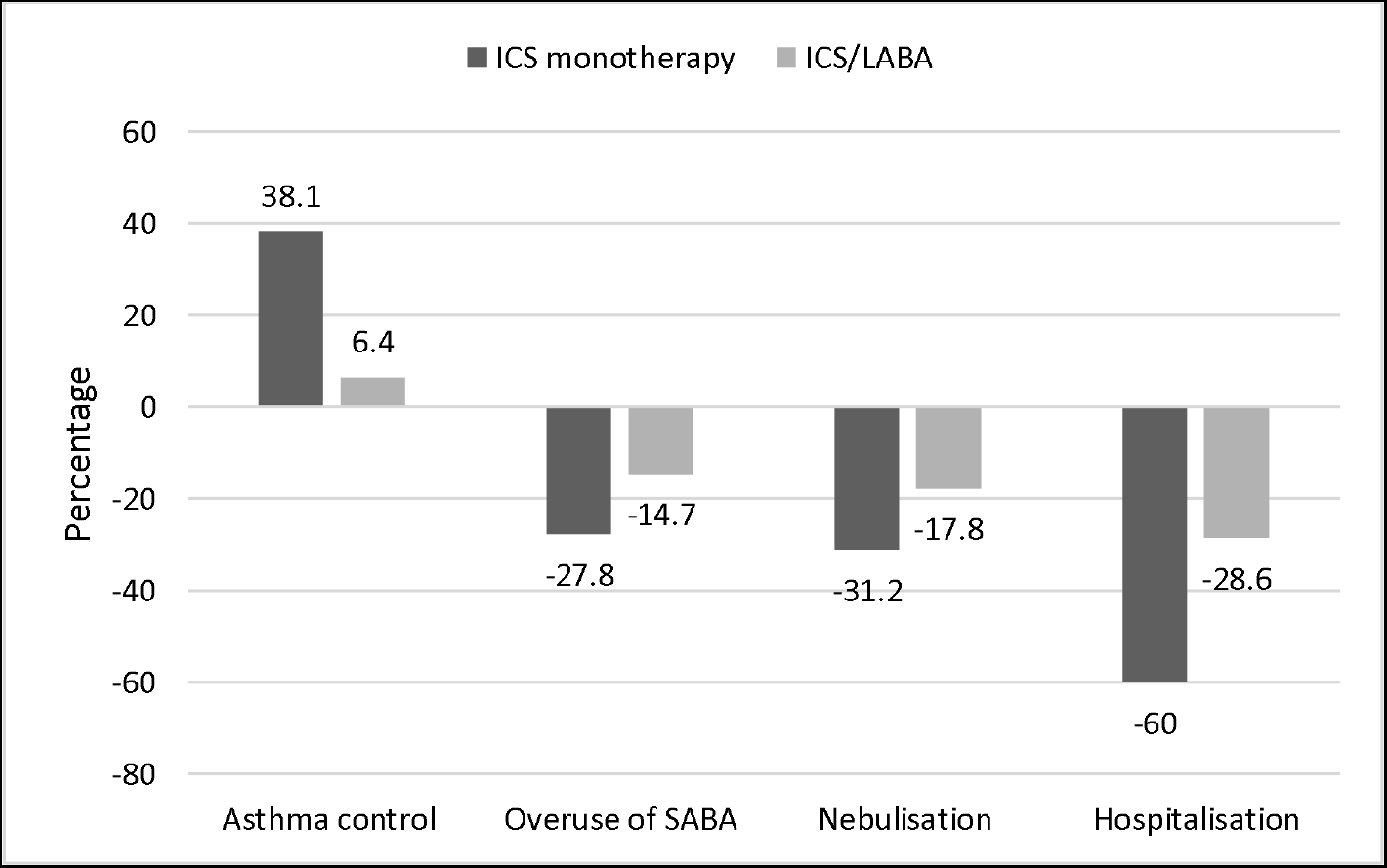
Percent changes in outcome measures. ICS-inhaled corticosteroid, LABA-long-acting beta_2_-agonist, SABA-short-acting beta_2_-agonist

When comparing baseline and second follow-up, a significant reduction in number of patients who forgot to take the therapy was observed in both ICS monotherapy (from 32.6% to 10.9%; p<0.001) and ICS/LABA (from 34.8% to 22.5%; p=0.002). However, reduction in number of patients who stopped the medications when they felt better did not show significant reduction in either group (monotherapy with ICS-from 81.3% to 77.6%; ICS/LABA - from 56.9% to 58.8%).

## Discussion

This study has generated evidence on the effectiveness of ICS monotherapy and ICS/LABA as opposed to many interventional studies which have documented the efficacy of these therapies. Effectiveness relates to how well a treatment works in real-life non-ideal circumstances, as opposed to efficacy, which measures how well it works in randomized controlled trial (RCT), or laboratory studies.[26] In fact, high-quality post-marketing observational studies are now considered as very important complement to the results of RCT in providing evidence on safety and effectiveness.[27]

This cohort study demonstrated that both ICS monotherapy and ICS/LABA were effective in controlling asthma. The age and gender distribution of our cohort (60% over 60 years, female preponderance) reflects the demography of the country.[25] Hence, our data could be generalizable to the country. However, there can be some inter-regional variation depending on other determinants of effectiveness such as appropriate use of inhaled medications, adherence and inhalation technique.[28]

In our cohort only about one-fourth (24.4%) of the participants were on ICS/LABA. This figure was very much lower than high-income counties [18, 29-30] but, it was higher than the figures reported from LMICs such as Kyrgyzstan, Nepal, Peru and Uganda. [9-11, 31]

A survey in Sri Lanka reported that the availability of ICS monotherapy in the public and private sectors was 81% and 75% respectively whereas the availability of ICS/LABA in the public and private sector was 17% and 63% respectively. [16] Since our cohort comprises patients receiving treatment in a public hospital, the low figure of 25% being on ICS/LABA is not unexpected. Poor economic access in the private sector could be the main cause for minimal use of ICS/LABA in this cohort of patients receiving treatment from public sector. Furthermore, we noticed that (Table 1) a greater proportion (63%) of participants on ICS monotherapy had low household income compared to those on ICS/LABA (41%). This strengthens our argument that poor economic access is the main cause of minimal use of ICS/LABA. In addition, we noticed that distribution of other sociodemographic factors such as age, sex, educational level, employment status and smoking showed comparable trends in both categories. These observations confirm access, as opposed to evidence and guidelines, to determine the type of inhaled medication for asthma in the present study.

We observed a progressive improvement in asthma control during the six-month follow-up in both groups. The proportion of patients with controlled asthma at the time of recruitment, first follow-up and second follow-up among those receiving ICS monotherapy were 54%, 66% and 72% respectively and that of participants on ICS/LABA were 76%, 79.4% and 80.9% respectively. This has been observed in other asthma cohort studies conducted in Turkey (61.5%, 82%, 84.8% and 87.3% in consecutive four visits) [32] and Poland (24.8%, 41.4%, 55.2% and 67.7% in consecutive four visits).[33] In our study percentage change in asthma control from baseline to second follow-up was greater (31.8%) in ICS monotherapy compared to ICS/LABA (6.4%). Further, a significant improvement in missed doses was observed from baseline to second follow-up in both groups. Interestingly improvement in asthma control in our cohort occurred without any change in the treatment of asthma. This brings in an important component in the treatment of asthma, close follow-up which ensures adherence.

It has been documented that non-adherence to medications is an important cause for poor asthma control.[34-35] Factors contributing to non-adherence include low perceived need for asthma medications, inadequate communication between patients and physicians, perceived concern regarding medications and inadequate knowledge.[20,31,35-36] Studies have also reported that close monitoring, frequent interactions with healthcare team and repeated instructions improve adherence to medications.[24,37-38] Patient education improves the patients’ understanding and adherence resulting in better asthma control.[39-40] All these can be addressed by a structured patient education programme which includes close monitoring, frequent interactions with healthcare team, providing basic facts about treatment, reassurance regarding side effects and repeated instructions. Although there was no formal patient education programme in our study plan, participants were free to communicate with the investigators during the follow-up. This could have given opportunity to the participants to clarify their concerns regarding the treatment which could have contributed to the improvement in adherence leading to improved asthma control. However, there was no significant change in the proportion of participants who stopped the treatment when they felt better in either group. This indicates inadequate knowledge of disease among the participants. As there was no formal patient education in our study, the impact on knowledge of disease among the participants would be less. This could explain why the proportion of participants who stopped asthma medication when they felt better remained high.

Traditionally, frequency of SABA use and number of acute attacks were the commonly used predictors for uncontrolled asthma.[19,29,41] We noticed that there was a significant reduction in overuse of SABA and number of nebulisations in ICS monotherapy group which was not seen in ICS/LABA group. In the local settings, because of the low availability, ICS/LABA is often reserved for difficult-to-control asthma. These patients are characterised by poor symptom control despite the regular treatment.[42] This could be the reason for lack of significant improvement in asthma control and predictors in those on ICS/LABA.

## Conclusions

Our findings have shown that both ICS monotherapy and ICS/LABA were effective in the treatment of asthma and improved over time. Non-pharmacological measures such as close follow-up and frequent communication with patients could have contributed to progressive improvement observed in this study. The impact of these measures was significant in those on ICS monotherapy but, limited in patients on ICS/LABA. Further, improved symptom control and reduction exacerbation would reduce the economic burden of asthma. Considering the low availability and affordability of ICS/LABA in the local settings, before switching to ICS/LABA, non-pharmacological measures such as regular follow-up, patient education and improving communication between patients and doctors must be tried in patients who fail to achieve adequate asthma control with ICS monotherapy. Inhaled corticosteroid/long-acting beta_2_-agonist could be offered to those who fail to achieve control with regular ICS monotherapy plus non-pharmacological measures. This approach would be more feasible and even cost-effective management strategy for asthma in Sri Lanka which is still struggling to recover from the recent economic crisis.

## Data Availability

All data produced in the present study are available upon reasonable request to the authors

## Acknowledgements

The authors would like to thank all participants who took part in the research and made this work possible. Mr. S. Thayapran, senior technical officer, Department of Pharmacology, Faculty of Medicine, University of Jaffna, Sri Lanka helped to enter the data in database.

## Contributors

YG, TSN, GS and SSR were involved in the research conception. Study was conceptualized by all four authors. YG was the principal investigator and responsible for the data collection, entry and analysis with TSN and SSR for manuscript preparation. All authors agree to be accountable for all aspects of the work. YG is the guarantor of this manuscript.

## Patient and public involvement

Patients and/or the public were not involved in the design, or conduct, or reporting, or dissemination of this study.

## Ethics approval and consent to participants

Approval for the study was obtained from the Ethics Review Committee, Faculty of Medicine, University of Colombo, Sri Lanka (EC-18-108). Permission was obtained from the Director, Teaching Hospital, Jaffna to collect the data. Written informed consent was obtained from all participants.

## Provenance and peer review

Not commissioned; externally peer-reviewed.

## Data availability statement

Data are available on reasonable request. The datasets used and/or analysed during the current study are available from the corresponding author on reasonable request.

## Open access

This is an open access article distributed in accordance with the Creative Commons Attribution Non Commercial (CC BY-NC 4.0) license, which permits others to distribute, remix, adapt, build upon this work non-commercially, and license their derivative works on different terms, provided the original work is properly cited, appropriate credit is given, any changes made indicated, and the use is non-commercial. See: http://creativecommons.org/licenses/by-nc/4.0/.

